# Meta-analysis of studies on the association between physician characteristics and patient outcomes

**DOI:** 10.1101/2025.01.22.25320956

**Authors:** Alyson Haslam, Sarah L. Miller, Vinay Prasad

## Abstract

**Objective:** Physicians have a large role in patient outcomes, and some have questioned whether some physician characteristics lead to better outcomes for the patient. Multiple studies have been conducted on the topic, leading to a variety of conclusions. To determine how frequently studies find favorable patient outcomes, by physician characteristics and the pooled estimated effect.

**Methods:** By searching PubMed, Web of Science, and Google Scholar, we identified studies that reported the association between physician characteristics (age, experience, sex/gender, race/ethnicity, nationality, and patient/physician characteristic concordance) and patient outcomes (patient outcomes, such as survival/mortality, hospitalization, readmissions, length of stay, and morbidity). We calculated the frequency of studies reporting on each type of physician characteristics and each type of patient outcome, risk of bias in included studies, and pooled estimates of association between physician characteristics and patient outcomes.

**Results:** Of the 32 included studies, the most common outcome was mortality. There were no differences in outcomes for physicians with more experience, compared with less experience (pooled odds ratio [OR]: 0.99; 95% confidence interval: 0.98 to 1.01; I^2^: 56.5%), between older and younger physicians (pooled OR: 1.01; 95% confidence interval: 0.97 to 1.05; I^2^: 68.1%), but there were significantly better outcomes for patients with a female physician, compared to patients with a male physician, although this difference was small and biased (pooled OR: 0.97; 95% confidence interval: 0.96 to 0.98; *I*^2^: 35.5%). Selection bias, unaccounted measurement in physician and patient characteristics, and not accounting for unintended exposures limit the findings of these types of studies.

**Conclusion:** Strategies to improve patient outcomes should focus on other factors. Moreover, when seeking care, patients should be more concerned about physician track record than non-modifiable characteristics.

## Introduction

The increasing costs of healthcare has necessitated a greater need in identifying factors associated with better value of care and improved patient outcomes. This is especially true in the US, where healthcare costs are about twice that of other high-income countries, yet life expectancy and infant mortality are often worse in the US.^1^

Physicians not only have a direct role in healthcare spending, through ordered tests and procedures,^2^ but also have an indirect role in the quality of care that patients receive, which has been associated with lower mortality.^3^ Because of their role in costs and outcomes, there has been an interest in examining physician characteristics in context of these outcomes. While some of these characteristics (e.g., communication style, work schedules, etc.) may be learned and developed, others (e.g., race, ethnicity, nationality, sex, and gender) may be considered immutable or purely academic because physicians may have little to no control over these characteristics.

Numerous studies have reported somewhat conflicting results on whether there is any association between physician characteristics and patient outcomes. Further, because characteristics such as race, ethnicity, sex, and gender cannot be randomized, studies examining these characteristics have had to rely on observational data, which can lead to biased outcomes. To our knowledge, there has not been a systematic evaluation of the totality of evidence, including an examination of the quality of these studies. As such, we sought to appraise the literature, systematically and comprehensively, for studies testing the association between physician characteristics and health outcomes.

## Methods

We sought to assess associations between physician characteristics (demographics, such as age, race, ethnicity, gender, sex, and/or nationality) and patient outcomes, including mortality, hospitalization, and morbidity.

### Search strategy

We searched PubMed, Google Scholar, and Web of Science using the search terms (doctor OR physician) AND (characteristics OR race OR gender OR sex OR ethnicity OR nationality OR sociodemographic) AND "patient outcomes". Our search date was June 16, 2023. We used the Publish or Perish Software to pull the first 1,000 search results from Google Scholar, but because of issues, the software was only able to pull the first 990 results.

While many important patient outcomes could have been considered, we limited patient outcomes to those relating to not only those that are most relevant to the patient, but are also more objective measures, such as hospitalizations, including readmissions and length of hospital stay, morbidity, or mortality. While we did not have a strict definition for morbidity, studies with a morbidity outcome needed to have specifically used “morbidity” as an outcome, as defined by the study authors.

We included original research studies that included physicians, examined the effect of physician demographic (age, race, ethnicity, gender, sex, nationality, and/or); and reported a patient outcome (hospitalizations (length of stay and readmissions), morbidity, and/or mortality) and the unit of analysis was the patient or physician. We included most original research studies, but we excluded search results if they were replies or commentaries to research articles, primary results of randomized studies (cannot randomize on these characteristics), pooled analyses, review articles, ecologic studies, and animal studies. We also excluded studies on non-demographic characteristics, such as education, training, patient load, experience, and communication style.

For each study, we noted information on characteristics, outcomes, sample size of patients and physicians, demographics of patient population, data source, years of observation, inclusion and exclusion criteria for patients and physician, and results, including odds ratios, uncertainty estimates, number of events, and number of admissions/surgeries. If studies reported multiple outcomes separately and as a composite, we abstracted data only on the individual outcomes so they were not being counted twice.

We used the adjusted results if available, but if adjusted point estimates and 95% confidence intervals were not provided, we used the numbers and percentages to calculate an unadjusted odds ratio and corresponding confidence intervals. We used the most aggregated data presented when studies presented both overall and stratified results. We combined studies reporting on race or ethnicity into the same category, and we combined the results from studies reporting on sex or gender into another category.

### Risk of bias assessment

For the risk of bias assessment, we based our questions on the AHRQ Risk of Bias questionnaire. Because our studies focused on non-interventional observational studies, we removed questions that were less relevant to the studies, and we added and modified questions that were not captured in the questionnaire but were relevant to this type of study. Specifically, we separated the question on confounding into 3 questions to ask about confounding and matching regarding patients, physicians, and hospitals. We added a question about using falsification endpoints. We also changed the question asking about pre-specified endpoints to ask about thoroughness of reporting the results. These questions were developed *a priori*, and responses were coded by two blinded reviewers. Risk of bias assessment was done independently by two reviewers blinded to each other’s responses (AH and SM). Discrepancies were discussed and reconciled by consensus.

### Statistical analysis

Data for this meta-analysis study were collected and analyzed using Excel, version 16.2 (Microsoft Corporation) and R statistical Software, version 4.2.1. We calculated study-level descriptive statistics. We used a Fleiss’ Kappa (R package ‘irr’) for determining agreement in risk of bias scores.

For each main characteristic (age, sex/gender, race/ethnicity, experience), we used the meta package of R to calculate pooled effect sizes, stratified by the major outcome (mortality, morbidity, hospitalization). For studies only reporting unadjusted frequencies of the outcome, we calculated an unadjusted odds ratio and corresponding 95% confidence intervals. Odds ratios for survival were recalculated so that they reflected the odds of death or mortality instead. We recalculated odds ratios so that they reflected the odds of outcome for females compared to males, US graduates compared to non-US, older and more years of experience compared to younger and fewer years of experience. We anticipated between-study heterogeneity, so we used a random effects model using the restricted maximum-likelihood estimator with Knapp-Hartung adjustments.

Our study involved publicly available data and did not involve individual patient data; thus, it was not submitted for institutional review board, in accordance with 45 CFR §46.102(f). We adhered to the Preferred Reporting Items for Systematic reviews and Meta-Analyses (PRISMA) reporting guidelines.

## Results

We found 4440 titles on PubMed (n=2247), Web of Science (n=1203), and Google Scholar (n=990), and after removing duplicates, we reviewed 3565 titles (Supplemental figure 1). After excluding studies not meeting our inclusion criteria, and including a study found during article review, 32 studies were left for inclusion and data analysis.^4–30^

The characteristics of the studies are presented in Table 1. The median patient sample size was 162444 (IQR: 25417, 725908; n=30), and the median physician sample size was 3314 (IQR: 299, 23390; n=23). The median age of participants was 62.0 (IQR: 52.0, 74.0; n=21), and the median percent of female participants was 51.8 (IQR: 42.4, 59.8; n=23).

**Table 1.** Characteristics of studies evaluating mortality, morbidity, and hospitalizations in relation to physician characteristics (N=32).

| Characteristic | Median (IQR) or n (%) |
| --- | --- |
| **Physician sample size, median (IQR; 23 studies)** | 3314 (299, 23390) |
| **Patient sample size, median (IQR, 30 studies)** | 162444 (25417, 725908) |
| **Patient age, median (IQR; 21 studies)** | 62.0 (52.0, 74.0) |
| **% of patients who are female, median (IQR; 23 studies)** | 51.8 (42.4, 59.8) |
| **Country, n (%)** |  |
| Canada | 5 (15.6) |
| France | 1 (3.1) |
| Japan | 1 (3.1) |
| Southeast Asia | 1 (3.1) |
| US | 24 (75.0) |
| **Study design, n (%)** |  |
| Case control | 1 (3.1) |
| Prospective (secondary analysis of RCT) | 1 (3.1) |
| Retrospective cross-sectional study | 2 (6.4) |
| Retrospective observational | 27 (84.4) |
| Survey | 1 (3.1) |
| **Physician data, n (%)** |  |
| American Hospital Association | 1 (3.1) |
| Community Tracking Study (CTS) Physician Survey | 1 (3.1) |
| Corporate Provider Database | 1 (3.1) |
| Doximity | 3 (9.4) |
| Florida Physician Workforce Survey | 2 (6.2) |
| Hospital/Medicare records | 7 (21.9) |
| Medical association master file | 7 (21.9) |
| Patient report | 1 (3.1) |
| Registry (same as patient) | 1 (3.1) |
| State/provincial database | 7 (21.9) |
| Trial data | 1 (3.1) |
| **Patient data, n (%)** |  |
| Hospital records | 4 (12.5) |
| Medicare | 9 (28.1) |
| Registry | 2 (6.2) |
| State/provincial database | 15 (46.9) |
| Survey | 1 (3.1) |
| Trial data | 1 (3.1) |
| **Physician type, n (%)** |  |
| Emergency | 6 (18.8) |
| General/internal medicine | 5 (15.6) |
| Hospitalist | 1 (3.1) |
| Multiple types included | 2 (6.2) |
| Nephrologist | 2 (6.2) |
| Not specified | 4 (12.5) |
| Surgery | 11 (34.4) |
| **Patient population, n (%)** |  |
| Condition | 13 (40.6) |
| Emergency | 1 (3.1) |
| General | 4 (12.5) |
| Hospitalization | 5 (15.6) |
| Surgery | 5 (15.6) |
| Not specified | 4 (12.5) |
| **Number of years of observation, median (IQR)** | 4.0 (2.0, 7.0) |
| **Year of publication, n (%)** |  |
| 1991 | 1 (3.1) |
| 1995 | 1 (3.1) |
| 2000 | 1 (3.1) |
| 2006 | 1 (3.1) |
| 2007 | 1 (3.1) |
| 2011 | 2 (6.2) |
| 2013 | 2 (6.2) |
| 2014 | 1 (3.1) |
| 2015 | 1 (3.1) |
| 2017 | 6 (18.8) |
| 2018 | 4 (12.5) |
| 2019 | 1 (3.1) |
| 2020 | 2 (6.2) |
| 2021 | 1 (3.1) |
| 2022 | 5 (15.6) |
| 2023 | 2 (6.2) |

Most study data involved a retrospective analysis of a cohort (27; 84.4%) and included patients and physicians from the US (24; 75.0%). Physician data were most commonly drawn from medical association master files (7; 21.9%) and hospital/Medicare records (7; 21.9%), whereas patient data most commonly came from state/provincial databases (15; 46.9%) and Medicare files (9; 28.1%). Patients were most often included based on a condition (13; 40.6%), but also based on if they were hospitalized (5; 15.6%) or for surgery (5; 15.6%). The median number of years of observation was 4.0 (IQR: 2.0,7.0).

There were 205 physician characteristic comparisons – 125 (61.0%) evaluated mortality, 51 (24.9%) evaluated length of stay, 13 (6.3%) evaluated morbidity, 11 (5.4%) evaluated readmission, 3 (1.5%) evaluated hospitalization, and 2 (1.0%) evaluated a composite endpoint.

Of the 205 comparisons, 73 (35.6%) were for experience, 62 (30.2%) were for age, 61 (29.8%) were for sex/gender, 2 (1.0%) were for nationality, and 7 (3.4%) were for race/ethnicity. Supplemental figure 3 shows the number of comparisons examining outcome type, by physician characteristic.

One study reported outcomes for 3 characteristics, 2 outcomes, in 16 conditions, but because of inadequacy in outcome reporting, we excluded this study, in addition to 2 other studies, for the pooled analyses.

### Risk of bias

The kappa statistic for agreement on risk of bias components was 0.68, which indicates moderate agreement. Overall, studies were mixed in their risk of bias. Studies were good at using uniform criteria for both comparison arms and using valid and reliable information for exposures, outcomes, and confounders, but many studies failed to fully adjust for confounding factors or limit unintended exposures, patients were able to self-select their provider, or there was an unclear description of missing information or missing information was high. Further, no study used a falsification endpoint to assess possible effects of confounding. Results indicating the percentage of studies meeting or not meeting risk of bias metrics are show in Supplemental Table 3.

### Experience

There were 9 studies (73 comparisons) for physician experience (Figure 1) – of which 19 (26.0%) found differences in outcomes with physician experience. In the pooled analysis (Figure 2), there was no difference in outcomes for physicians with more experience, compared with less experience (pooled odds ratio [OR]: 0.99; 95% confidence interval: 0.98 to 1.01; *I*^2^: 56.5%), but there were significant differences between outcome type (p<0.0001). There was no difference in mortality (1.00; 95% confidence interval: 0.98 to 1.01; *I*^2^: 54.6%; 26 comparisons) or morbidity (0.96; 95% confidence interval: 0.76 to 1.21; *I*^2^: 72.6%; 6 comparisons), but patients with less experienced physicians had lower odds of a readmission, compared to patients with more experienced physicians (0.92; 95% confidence interval: 0.90 to 0.95; *I*^2^: 0.0%; 6 comparisons).

**Figure 1.**
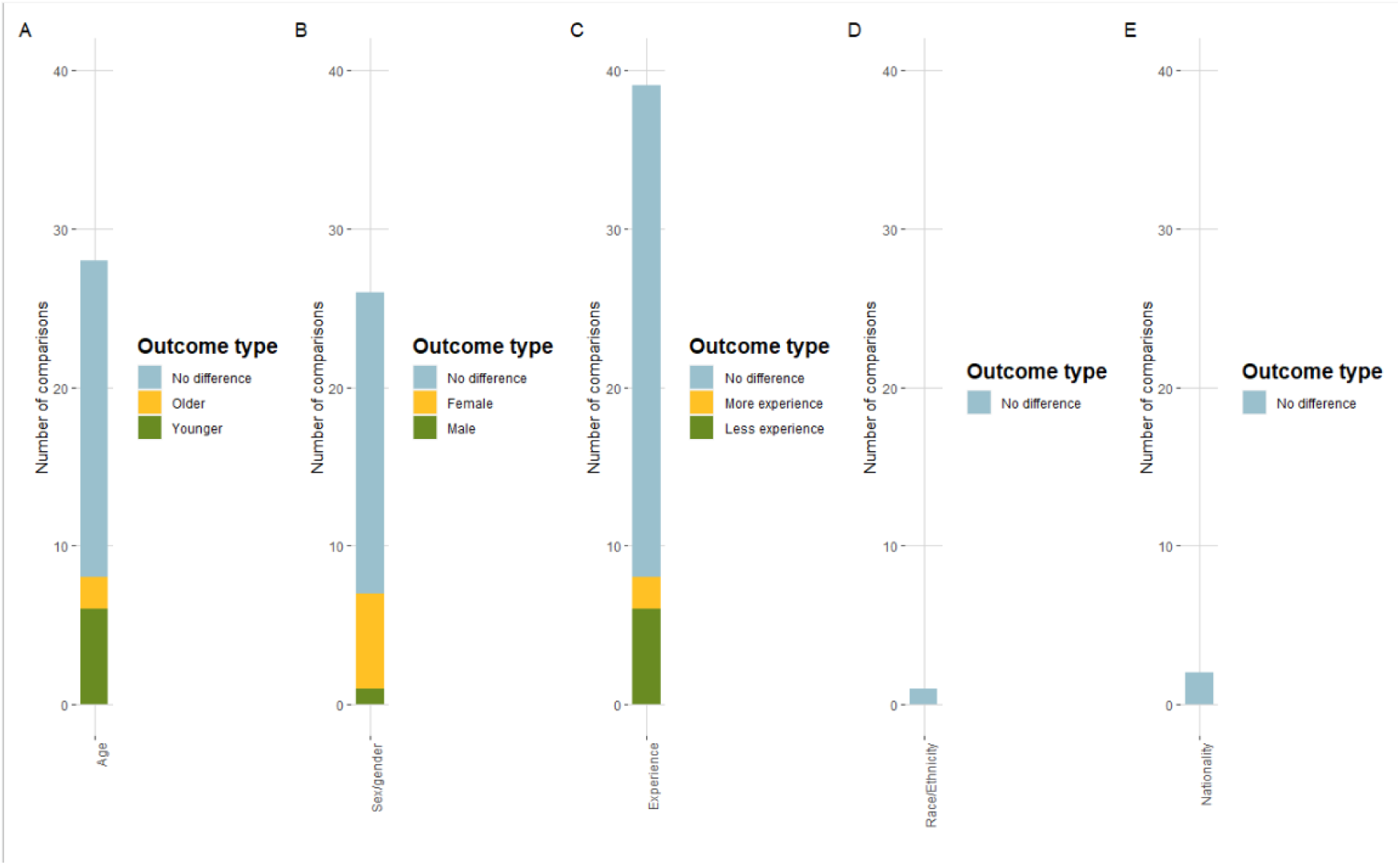
Number of comparisons showing favorable patient outcomes, by physician characteristic, in studies examining the effect of physician characteristics on patient outcomes. A=age, B=sex/gender, C=experience, D=Race/ethnicity, and E=nationality.

**Figure 2.**
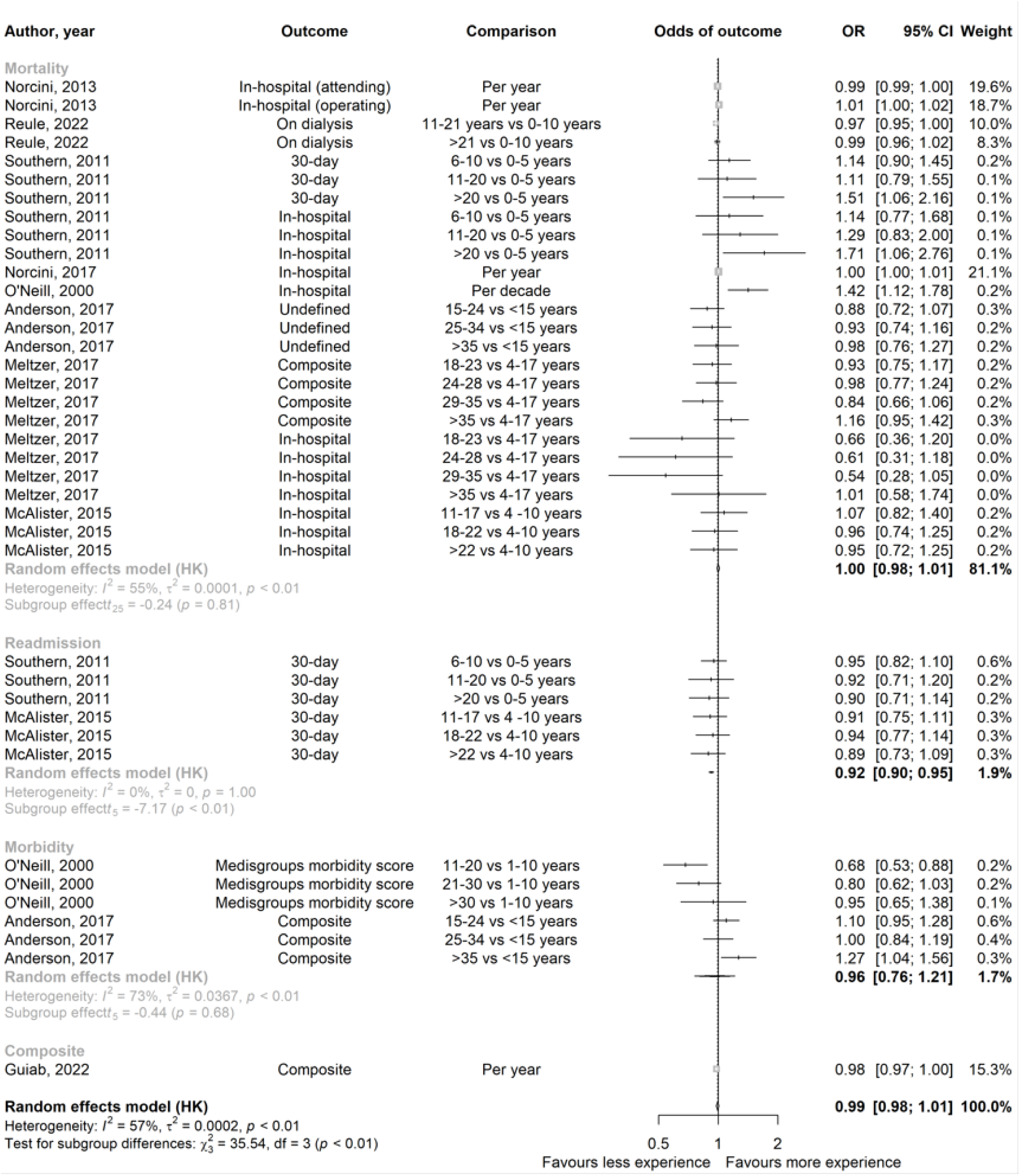
Pooled odds of patient outcomes for studies assessing physician experience.

### Age

There were 13 studies (62 comparisons) for physician age (Figure 1) – of which 33 (53.2%) found differences in outcomes with physician age. In the pooled analysis (Figure 3), there were no differences in patient outcomes between older and younger physicians (pooled OR: 1.01; 95% confidence interval: 0.97 to 1.05; *I*^2^: 68.1%), and there were no significant differences between outcome type (p=0.07). There were no differences in mortality (1.02; 95% confidence interval: 0.98 to 1.07; *I*^2^: 69.8%; 24 comparisons), morbidity (0.84; 95% confidence interval: 0.59 to 1.21; *I*^2^: 0.0%; 3 comparisons), or readmission (1.00; 95% confidence interval: 0.99 to 1.01; *I*^2^: 0.0%; 1 comparison).

**Figure 3.**
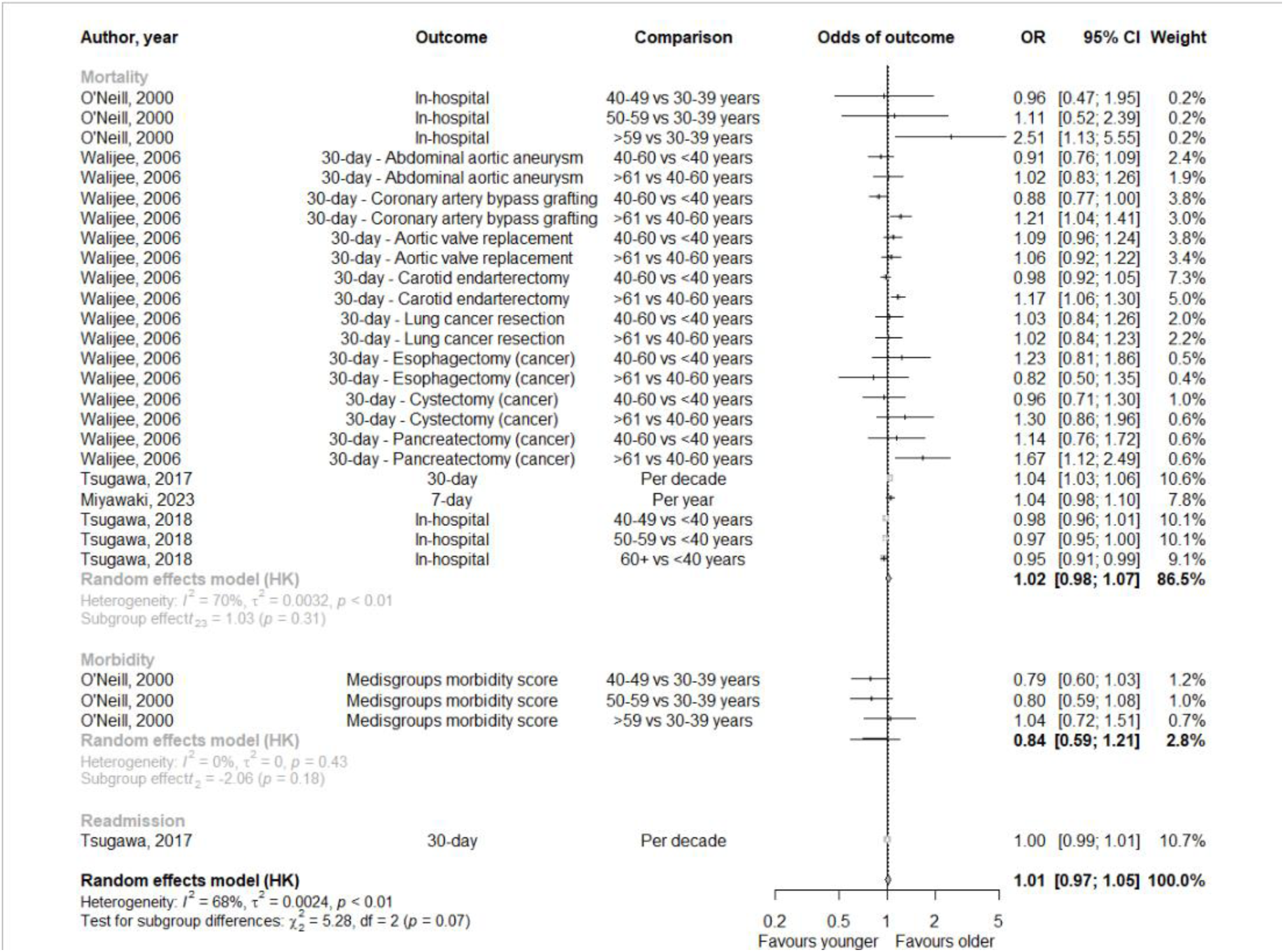
Pooled odds of patient outcomes for studies assessing physician age.

### Sex/gender

There were 14 studies (61 comparisons) for physician sex/gender (Figure 1) – of which 19 (31.1%) found differences in patient outcomes with physician sex/gender. In the pooled analysis (Figure 4), patients with a female physician had lower odds of an adverse patient outcome, compared to patients with a male physician (pooled OR: 0.97; 95% confidence interval: 0.96 to 0.98; *I*^2^: 35.5%), but there were significant differences between outcome type (p=0.04). There was no difference in hospitalization (0.96; 95% confidence interval: 0.83 to 1.11; *I*^2^: 58.8%; 3 comparisons), morbidity (1.07; 95% confidence interval: 0.45 to 2.51; *I*^2^: 0.0%; 2 comparisons), or readmission (0.97; 95% confidence interval: 0.91 to 1.03; *I*^2^: 0.0%; 2 comparisons), but patients with a female physician had lower odds of mortality (0.96; 95% confidence interval: 0.95 to 0.97; *I*^2^: 40.3%; 15 comparisons) and length of stay (0.98; 95% confidence interval: 0.96 to 0.99; *I*^2^: 0.0%; 3 comparisons).

**Figure 4.**
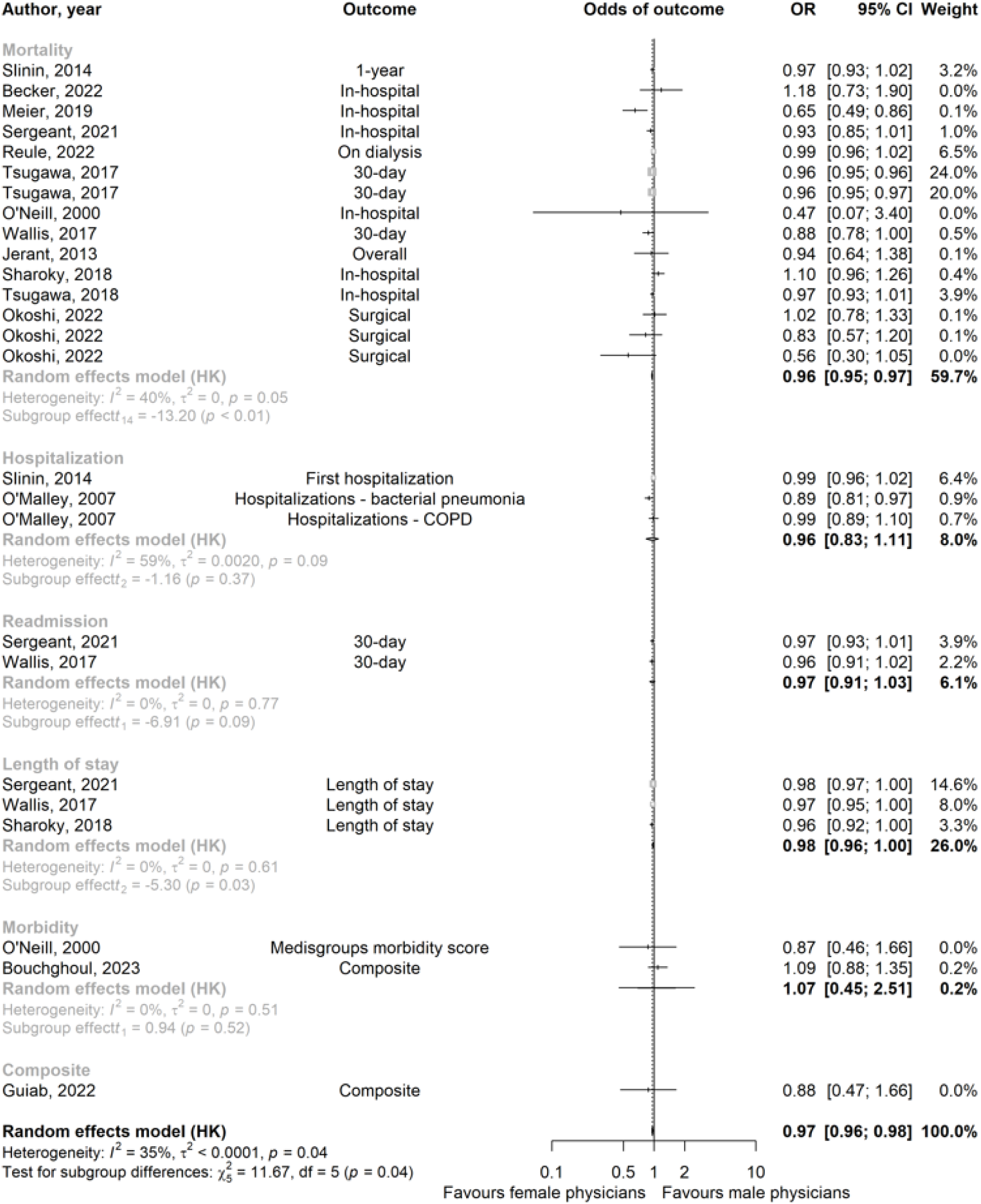
Pooled odds of patient outcomes for studies assessing physician sex/gender.

### Race/ethnicity and nationality

There were six studies (7 comparisons) that evaluated race/ethnicity. Four comparisons examined racial concordance between patient and physicians instead of specific patterns by race. Two comparisons lacked specific outcome data and just reported that there were no differences. For the one study that reported specific outcome data, there were no differences in mortality between whites and other race/ethnicities (OR: 0.86; 95% CI: 0.61 to 1.21). There was only one study that reported on nationality. There were no differences in mortality (OR: 1.01; 95% CI: 0.95 to 1.07) or readmission (OR: 1.00; 95% CI: 0.97 to 1.03) between US physicians and non-US physicians.

### Concordance

There were 4 studies that evaluated concordance (1 for sex/gender and 3 for race/ethnicity). In the pooled analysis (Supplemental figure 4), there were no differences in mortality or readmission (beta coefficient: -0.0005; 95% CI: -0.003 to 0.002; *I*^2^: 84.7%) by race/ethnicity, but the one study examining sex/gender concordance found lower mortality when patients and physicians were concordant (beta coefficient: -0.01; 95% CI: -0.01 to -0.007).

## Discussion

We found that physician characteristics, such as age, experience, race/ethnicity, and nationality, lack statistical association with patient mortality. There was a significant association between having a female physician and lower patient mortality, but the difference was very small. Overall, we noted that confounding could have a notable influence on the results of these studies.

One of the major limitations of these study types is that they are observational, and the exposures cannot be tested interventionally. Because they are observational, the findings from the studies are prone to many biases – biases that are unlikely to be fully accounted for in the analysis.

One way to test for unmeasured confounding is through falsification endpoints, which are endpoints that are highly unlikely to be related to the exposure (e.g., height or hemoglobin concentration of patient prior to physician interaction).^31^ When these endpoints are included in the analysis, the presence of significant associations between the falsification endpoint and exposure, may indicate the need for caution when interpreting study results. We noted that there were no studies that included this type of endpoint to evaluate confounding.

The characteristics being studied, in addition to being inherent and unchangeable, may be more likely to be a surrogate for other characteristics that are modifiable. For example, better outcomes for patients with female physicians may be due to the fact that women have historically been underrepresented in medicine.^32^ The few women who have practiced medicine may be more exceptional because they have had more obstacles to overcome, whereas male physicians may be more representative of males, in general, because there have been fewer obstacles to overcome to get into the medical field. Or, it may be that women physicians are more likely to be caring and compassionate, but these are characteristics that can be developed by anyone.

Many of our studies included surgeons, of which females make up only a small fraction of the specialty,^32^ yet only two studies ran their analysis on a matched cohort.^25,28^ The lack of statistical adjustment done through matching is likely to lead to unmeasured confounding when representation is so disproportionate.

### Limitations

Our study has several limitations. First, we reviewed references included in prior reviews on this topic, which may have led to more included articles for certain specialties. Hence, our findings may be less generalizable to the literature at-large. Second, we also may not have found all articles testing this association, but we searched 3 common databases using broad search terms and searched through about 3500 articles. Third,

## Conclusion

In conclusion, we found a lack of association between most physician characteristics and patient outcomes. Patients with female physicians have significantly better outcomes, but the effect size is very small. Strategies to improve patient outcomes, including mortality and hospitalizations, should focus on other modifiable factors with larger impacts.

## Data Availability

All data produced in the present study are available upon reasonable request to the authors

## Authorship contribution

VP and AH conceptualized study design; AH, and SM reviewed and abstracted data; VP reviewed and confirmed abstracted data; AH wrote first draft of manuscript; and VP and SM reviewed and revised subsequent and finalized draft of manuscript.

---

**Supplemental Figure 1:**
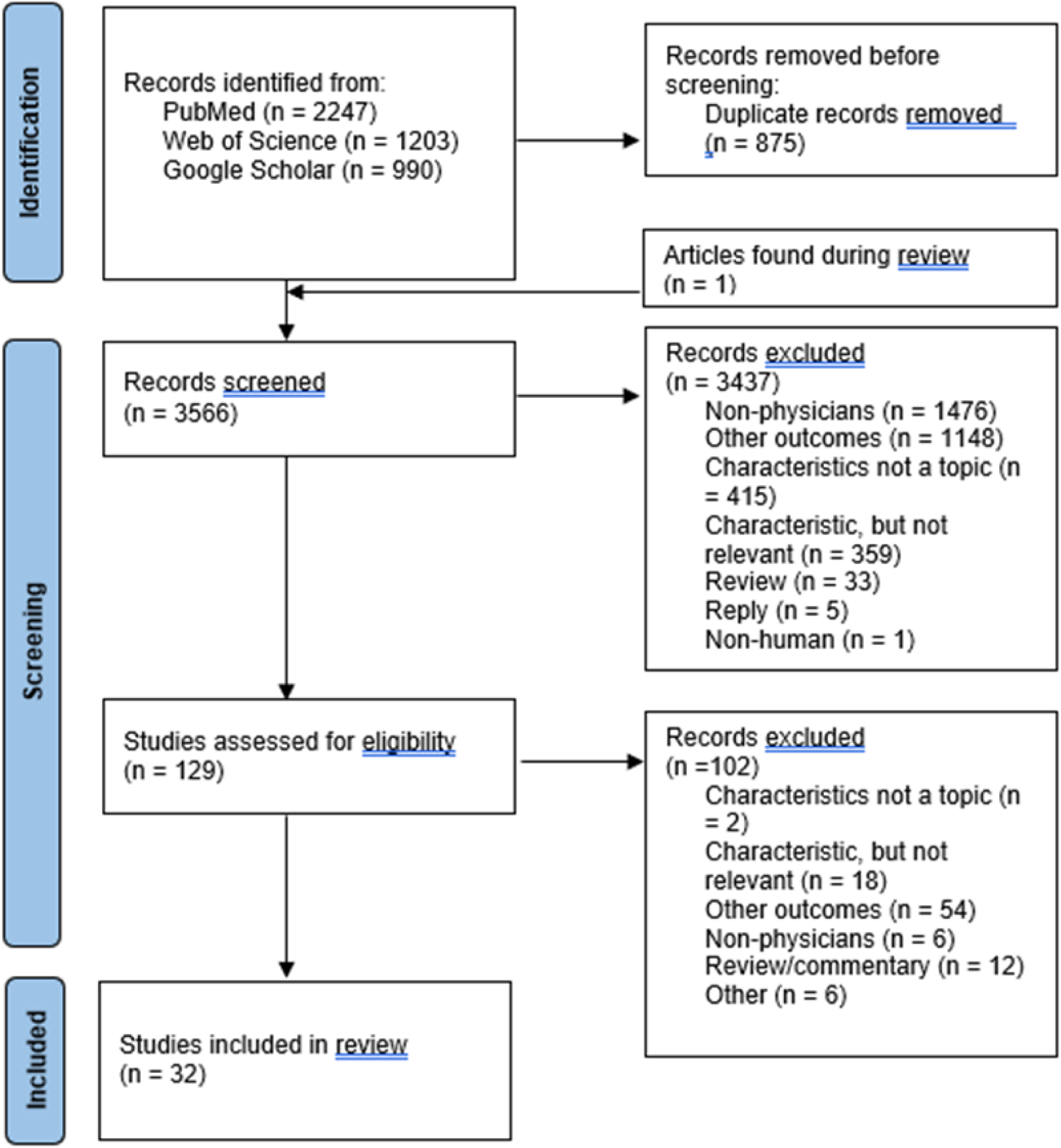
Flow chart detailing the inclusion of studies examining the association between physician characteristics and patient outcomes.

**Supplemental Figure 2.**
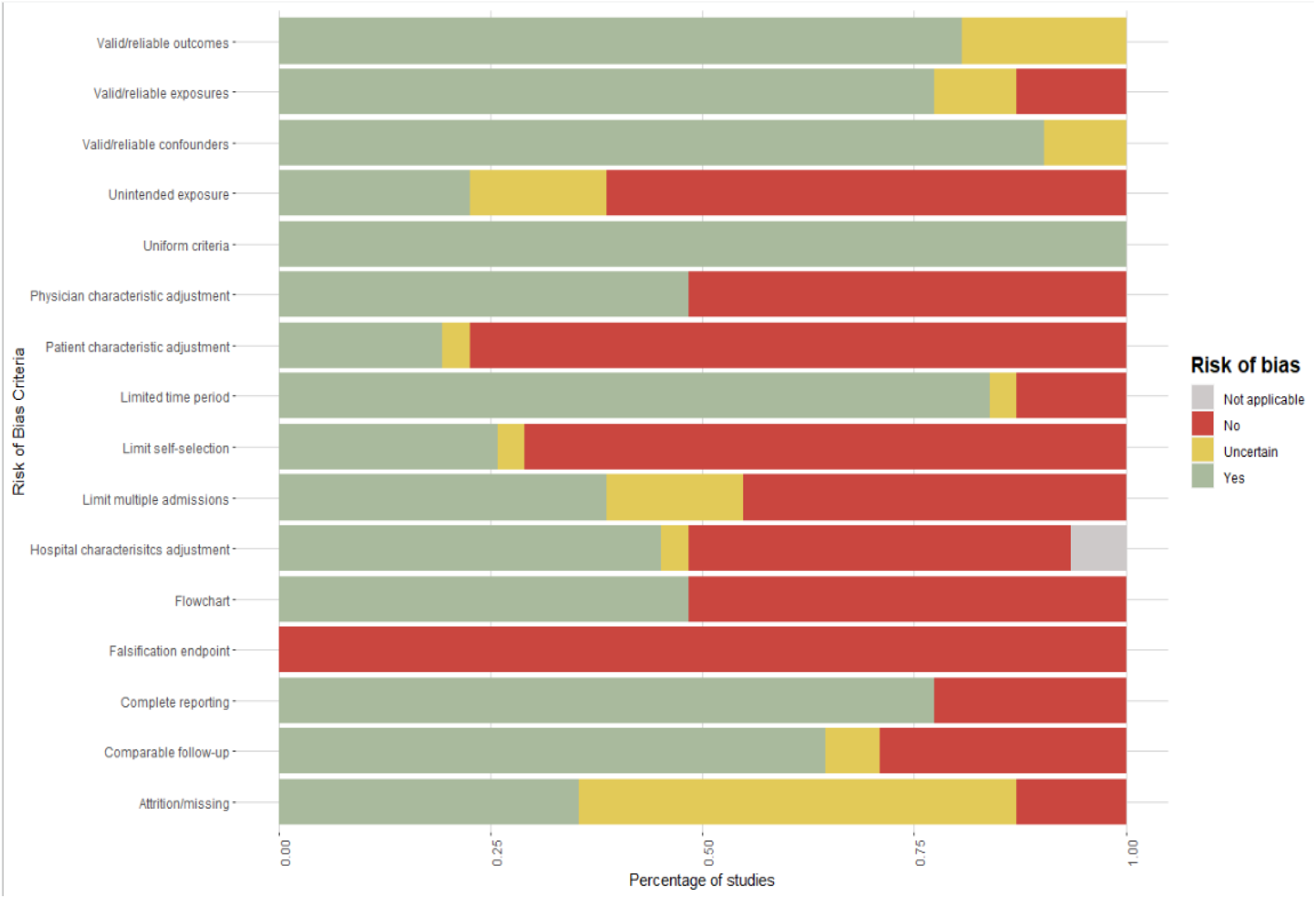
Risk of bias assessment for studies evaluating physician characteristics and patient outcomes

**Supplemental Figure 3.**
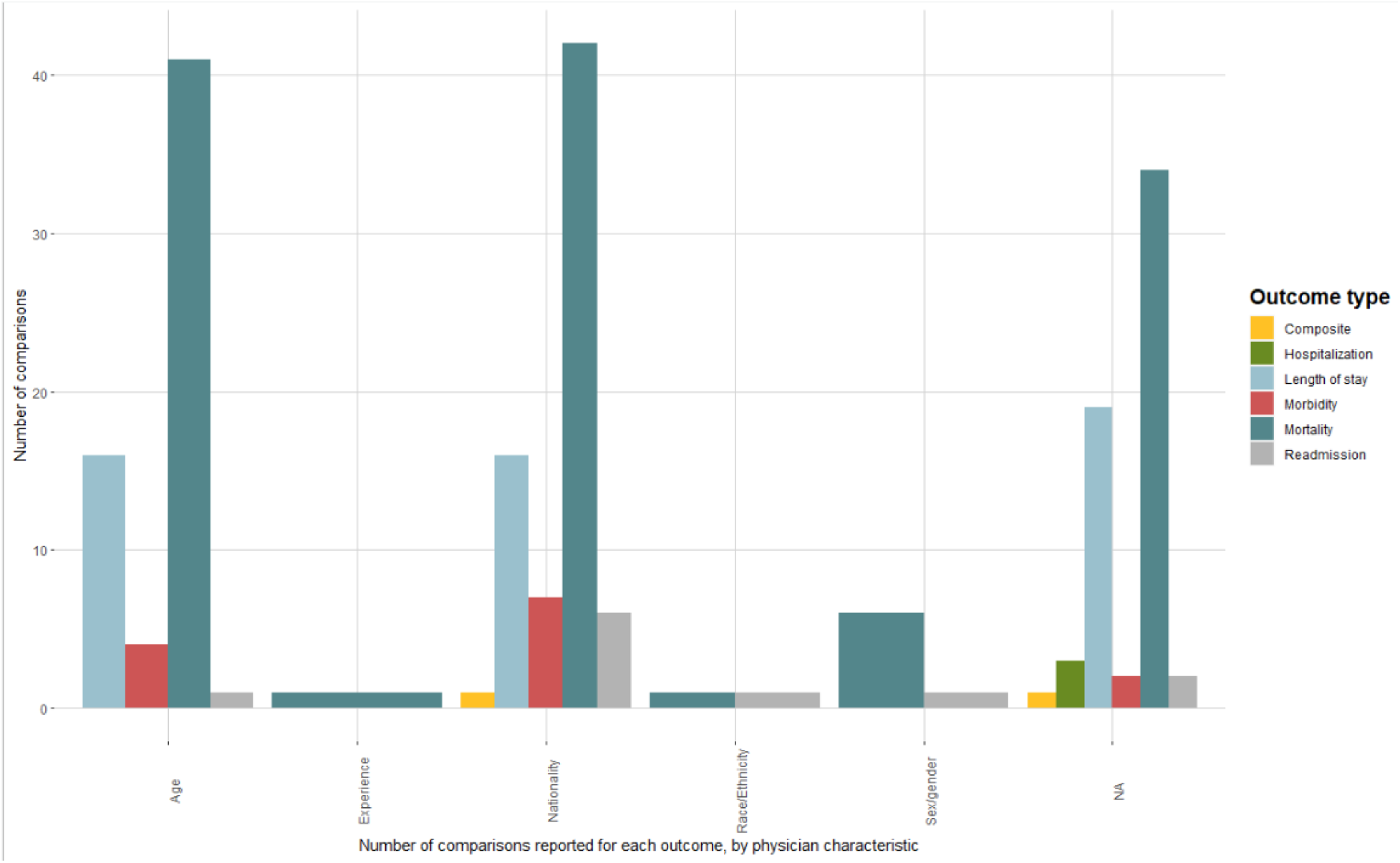
Number of comparisons examining outcome type, by physician characteristic.

**Supplemental Figure 4.**
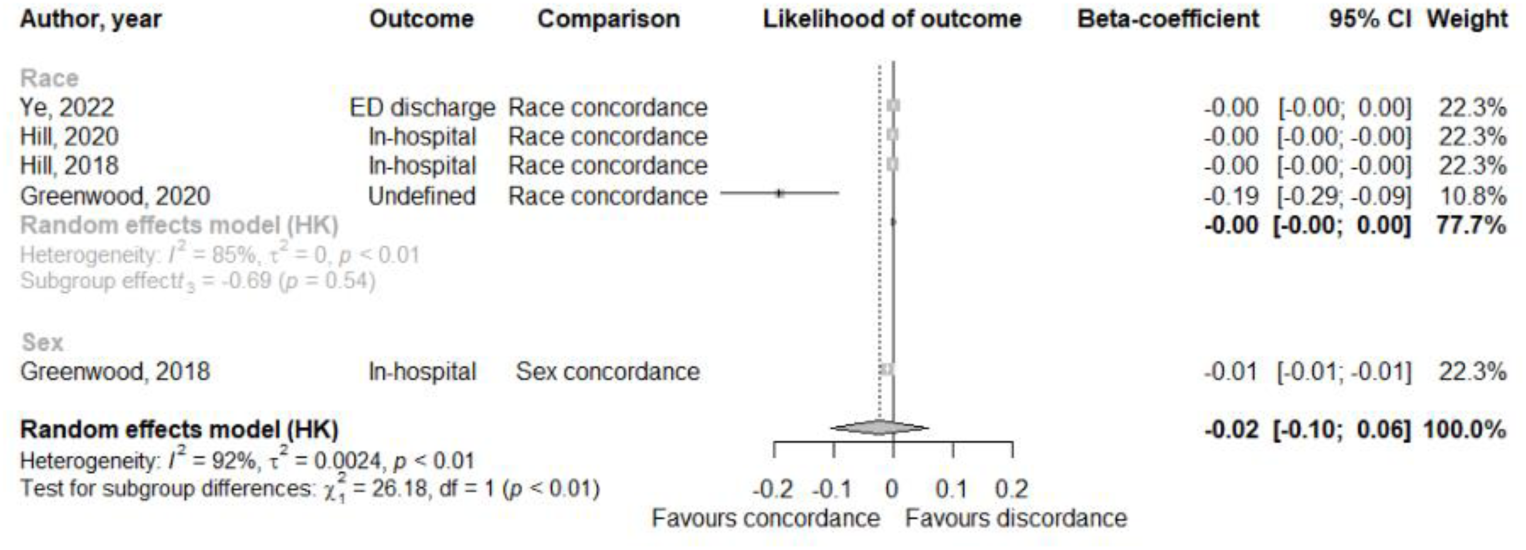
Pooled regression coefficients of patient outcomes for studies assessing physician-patient concordance, by physician characteristic.

## References

1. Papanicolas I, Woskie LR, Jha AK. Health Care Spending in the United States and Other High-Income Countries. JAMA. 2018;319(10):1024–1039. doi:10.1001/jama.2018.1150

2. Fred HL. Cutting the cost of health care: the physician’s role. Tex Heart Inst J. 2016;43(1):4–6. doi:10.14503/THIJ-15-5646

3. Pereira Gray DJ, Sidaway-Lee K, White E, Thorne A, Evans PH. Continuity of care with doctors-a matter of life and death? A systematic review of continuity of care and mortality. BMJ Open. 2018;8(6):e021161. doi:10.1136/bmjopen-2017-021161

4. Slinin Y, Guo H, Li S, et al. Hemodialysis patient outcomes: provider characteristics. Am J Nephrol. 2014;39(5):367–375. doi:10.1159/000362286

5. Becker L, Siry-Bove BJ, Shelton SK, McDaniel K, Nelson JL, Perman SM. Does Physician Gender and Gender Composition of Clinical Teams Affect Guideline Concordance and Patient Outcomes in Out-of-Hospital Cardiac Arrest? J Womens Health (Larchmt). 2022;31(12):1800–1804. doi:10.1089/jwh.2021.0399

6. Norcini JJ, Boulet JR, Opalek A, Dauphinee WD. Outcomes of cardiac surgery: associations with physician characteristics, institutional characteristics, and transfers of care. Med Care. 2013;51(12):1034–1039. doi:10.1097/MLR.0b013e3182a048af

7. Davidson W, Molloy DW, Bédard M. Physician characteristics and prescribing for elderly people in New Brunswick: relation to patient outcomes. CMAJ. 1995;152(8):1227–1234.

8. Meier A, Yang J, Liu J, et al. Female physician leadership during cardiopulmonary resuscitation is associated with improved patient outcomes. Crit Care Med. 2019;47(1):e8–e13. doi:10.1097/CCM.0000000000003464

9. Sergeant A, Saha S, Shin S, et al. Variations in processes of care and outcomes for hospitalized general medicine patients treated by female vs male physicians. JAMA Health Forum. 2021;2(7):e211615. doi:10.1001/jamahealthforum.2021.1615

10. Reule S, Foley R, Shaughnessy D, Drawz P, Ishani A, Rosenberg M. Does experience matter? The relationship between nephrologist characteristics and end stage kidney disease patient outcomes. Hemodial Int. 2022;26(1):114–123. doi:10.1111/hdi.12961

11. Southern WN, Bellin EY, Arnsten JH. Longer lengths of stay and higher risk of mortality among inpatients of physicians with more years in practice. Am J Med. 2011;124(9):868–874. doi:10.1016/j.amjmed.2011.04.011

12. Tsugawa Y, Jena AB, Figueroa JF, Orav EJ, Blumenthal DM, Jha AK. Comparison of hospital mortality and readmission rates for Medicare patients treated by male vs female physicians. JAMA Intern Med. 2017;177(2):206–213. doi:10.1001/jamainternmed.2016.7875

13. Tsugawa Y, Newhouse JP, Zaslavsky AM, Blumenthal DM, Jena AB. Physician age and outcomes in elderly patients in hospital in the US: observational study. BMJ. 2017;357:j1797. doi:10.1136/bmj.j1797

14. Tsugawa Y, Jena AB, Orav EJ, et al. Age and sex of surgeons and mortality of older surgical patients: observational study. BMJ. 2018;361:k1343. doi:10.1136/bmj.k1343

15. Miyawaki A, Jena AB, Burke LG, Figueroa JF, Tsugawa Y. Association between emergency physician’s age and mortality of medicare patients aged 65 to 89 years after emergency department visit. Ann Emerg Med. 2023;82(3):301–312. doi:10.1016/j.annemergmed.2023.02.010

16. Norcini JJ, Boulet JR, Opalek A, Dauphinee WD. Patients of doctors further from medical school graduation have poorer outcomes. Med Educ. 2017;51(5):480–489. doi:10.1111/medu.13276

17. O’Neill L, Lanska DJ, Hartz A. Surgeon characteristics associated with mortality and morbidity following carotid endarterectomy. Neurology. 2000;55(6):773–781. doi:10.1212/wnl.55.6.773

18. Anderson BR, Wallace AS, Hill KD, et al. Association of surgeon age and experience with congenital heart surgery outcomes. Circ Cardiovasc Qual Outcomes. 2017;10(7). doi:10.1161/CIRCOUTCOMES.117.003533

19. Waljee JF, Greenfield LJ, Dimick JB, Birkmeyer JD. Surgeon age and operative mortality in the United States. Ann Surg. 2006;244(3):353–362. doi:10.1097/01.sla.0000234803.11991.6d

20. Guiab K, Evans T, Brigode W, et al. Complications After Inpatient Laparoscopic Cholecystectomy: Effect of Surgeon Experience, Procedure Volume, and Other Surgeon-Based Characteristics. Am Surg. 2022;88(8):1798–1804. doi:10.1177/00031348221084083

21. Meltzer AJ, Agrusa C, Connolly PH, Schneider DB, Sedrakyan A. Impact of provider characteristics on outcomes of carotid endarterectomy for asymptomatic carotid stenosis in new york state. Ann Vasc Surg. 2017;45:56–61. doi:10.1016/j.avsg.2017.05.015

22. Burns LR, Wholey DR. The effects of patient, hospital, and physician characteristics on length of stay and mortality. Med Care. 1991;29(3):251–271. doi:10.1097/00005650-199103000-00007

23. Chidambaram M, Bargman JM, Quinn RR, Austin PC, Hux JE, Laupacis A. Patient and physician predictors of peritoneal dialysis technique failure: a population based, retrospective cohort study. Perit Dial Int. 2011;31(5):565–573. doi:10.3747/pdi.2010.00096

24. McAlister FA, Youngson E, Bakal JA, Holroyd-Leduc J, Kassam N. Physician experience and outcomes among patients admitted to general internal medicine teaching wards. CMAJ. 2015;187(14):1041–1048. doi:10.1503/cmaj.150316

25. Wallis CJ, Ravi B, Coburn N, Nam RK, Detsky AS, Satkunasivam R. Comparison of postoperative outcomes among patients treated by male and female surgeons: a population based matched cohort study. BMJ. 2017;359:j4366. doi:10.1136/bmj.j4366

26. O’Malley AS, Pham HH, Schrag D, Wu B, Bach PB. Potentially avoidable hospitalizations for COPD and pneumonia: the role of physician and practice characteristics. Med Care. 2007;45(6):562–570. doi:10.1097/MLR.0b013e3180408df8

27. Jerant A, Bertakis KD, Fenton JJ, Franks P. Gender of physician as the usual source of care and patient health care utilization and mortality. J Am Board Fam Med. 2013;26(2):138–148. doi:10.3122/jabfm.2013.02.120198

28. Sharoky CE, Sellers MM, Keele LJ, et al. Does surgeon sex matter?: practice patterns and outcomes of female and male surgeons. Ann Surg. 2018;267(6):1069–1076. doi:10.1097/SLA.0000000000002460

29. Bouchghoul H, Deneux-Tharaux C, Georget A, et al. Association between surgeon gender and maternal morbidity after cesarean delivery. JAMA Surg. 2023;158(3):273–281. doi:10.1001/jamasurg.2022.7063

30. Okoshi K, Endo H, Nomura S, et al. Comparison of short term surgical outcomes of male and female gastrointestinal surgeons in Japan: retrospective cohort study. BMJ. 2022;378:e070568. doi:10.1136/bmj-2022-070568

31. Prasad V, Jena AB. Prespecified falsification end points: can they validate true observational associations? JAMA. 2013;309(3):241–242. doi:10.1001/jama.2012.96867

32. AAMC. Active Physicians by Sex and Specialty, 2021. December 31, 2021. Accessed October 27, 2023. https://www.aamc.org/data-reports/workforce/data/active-physicians-sex-specialty-2021

